# “Lived Experiences, Impacts, and Coping Strategies During COVID-19: A Qualitative Study of Frontline Health Care Workers in a Zonal Hospital in Northern Tanzania”

**DOI:** 10.64898/2026.02.04.26345569

**Authors:** Lisbeth Mhando, Gudina Terefe Tucho, Tania Aase Draebel, Lena Skovgaard Andersen, Declare L. Mushi, Reginald Kavishe

## Abstract

**Background:** The COVID-19 pandemic significantly changed the daily routines of frontline health workers (FLHW), particularly those directly caring for COVID-19 patients. This study explores the lived experiences and coping strategies of FLHW at a Zonal Hospital in Kilimanjaro, Tanzania

**Method:** The study used a qualitative exploratory descriptive design, to retrospectively capture psychological impacts, challenges, coping strategies, and professional dilemmas faced by FLHW. Participants were purposively sampled from KCMC Hospital departments directly involved in COVID-19 patient care.

**Findings:** The FLHW experienced considerable fear, stress, and stigma. The fear of infection and high mortality rates among patients and colleagues heightened their anxiety. Stress was exacerbated by long working hours, uncomfortable personal protective equipment (PPE), poor remuneration, and the emotional toll of witnessing numerous deaths. FLHW also experienced being stigmatized and discriminated against, both within their workplaces, within the family and in the broader community. Some FLHW considered quitting their jobs due to the overwhelming workload, fear of being infected, and emotional strain. Additionally, absenteeism and avoidance of COVID-19 duties were common, driven by fear and inadequate hospital capacity. Coping strategies among FLHW to manage their stress and maintain resilience included acceptance, faith, family support, rest, and, the use of recreational substances.

**Conclusion:** The FLHW experienced considerable fear, stress, and stigma. The study highlights the need for better psychological support, improved communication, adequate training, and resources to support FLHW before, during, and post-pandemic. Enhanced resilience and confidence, along with a greater appreciation for protective measures and compassion for patients, were some of the key lessons learned from their experiences during the pandemic informing more effective preparedness care in future pandemics.

## Background

The pandemic of coronavirus disease 2019 (COVID-19) caused significant changes in the everyday routine of health workers, especially those in direct contact with COVID-19 patients [1,2]. The impact of this pandemic has been felt worldwide and as of September 2022, the World Health Organization had recorded 612,724,171 confirmed cases, including 6,517,123 deaths [3].

Frontline health workers (FLHW) spent long hours during the pandemic caring for patients and experienced drastic changes in their working environment leaving them at risk of developing mental health challenges that interfere with overall wellbeing. The sudden onset and magnitude of the disease also left many FLHW incapable of dealing with the demands and uncertainty of the progression of the disease [4].

Tanzania like other countries across the globe experienced challenges during the COVID-19 pandemic. The impact of COVID-19 on frontline health workers in Tanzania was profound affecting their physical and mental wellbeing as they struggled to maintain work-life balance in a limited resource setting. Although the pandemic is behind us now, the experience of COVID-19 was a unique learning point for health workers and government systems on the importance of pandemic preparedness [3]. In this study,’frontline healthcare workers’ refers to medical professionals directly involved in patient care, including doctors, nurses, and support staff. This paper explores the lived experiences of FLHW at a zonal hospital in Kilimanjaro region, Tanzania during the COVID-19 pandemic.

## Materials and methods

### Study design

A retrospective qualitative study was conducted from 24 /1/ 2024 to 20/2/ 2024 to capture the lived experiences of health workers and coping strategies who attended to patients with COVID-19 and their relatives. Ethical approval was granted by IRBs from Kilimanjaro Medical University College (KCMUCo) and the National Institute for Medical Research (NIMR)

### Study site and sample population

The study was conducted at Kilimanjaro Christian Medical Center (KCMC) Zonal Referral Hospital in Northern Tanzania which was of the zonal hospitals elected for treatment of COVID 19 cases. We involved Hospital managers and healthcare workers who had direct contact with patients undergoing treatment for COVID-19 and their relatives.

### Sample selection

Purposive sampling was employed to select 14 study participants from departments that were directly responsible for providing care to patients diagnosed with COVID-19 during the pandemic which were emergency, internal medicine (managed the isolation ward), and pathology departments. Participants included ambulance drivers, mortuary workers, doctors, nurses, and members of the COVID-19 prevention committee. Sample was determined by data saturation in the data collection process.

### Data collection tools and methods

An in-depth Interview guide was developed by the research team guided by objectives of the study and existing literature. The guide was developed in English and later translated to Kiswahili by a mental health expert. The translated guide was shared with another team member to assess for accuracy and clarity before it was used for data collection. The in-depth interviews were conducted to explore the study’s various themes such as the psychological impact of COVID-19, lived experiences of hospital workers, challenges and barriers to accessing psychological support, stress management, coping strategies, skills, and the institutional support received during the COVID-19 pandemic. Written consent was obtained from all participants prior Interview. In-depth interviews were conducted in Kiswahili language and audio recorded by two trained research assistants. Field notes were taken during the interview process and expanded within 24 hours after the interview. The researchers had backgrounds and experiences in the mental health field. Interviews were conducted in a quiet place in the hospital and they lasted for about 40 minutes.

### Data analysis and interpretation

Data was transcribed verbatim in the Kiswahili language and then translated into English. The research team read each transcript to achieve a clear understanding of the statements and gain familiarity with the data. Using thematic analysis, information with similar themes was grouped. Coding was then done by two members of the study team for inter-rater reliability. The codes developed from the transcript were used to identify patterns of meaning in the collected data, which were then used to create key emerging themes.

## Results

### Participants’ social demographic characteristics

The study involved 14 participants. Six (6) participants were male and eight (8) females. Eleven participants were married, single were 3, and divorced was 1. Five (5) medical doctors, 5 nurses, 2 administrative staff and 2 health care managers. The age of study participants ranged between 28 and 60 years, mean age was 42.4 years. A total of 10 participants had tested for COVID-19, six (6) were infected with COVID-19 and 10 were vaccinated. Detailed social demographic characteristics are presented in Table 1

**Table 1:** Social demographic characteristics of study participants (No.14)

| **Characteristics** |  | **No** |
| --- | --- | --- |
| Sex | Male | 6 |
|  | Female | 8 |
| Age | 20-35 | 5 |
|  | 36-50 | 8 |
|  | 51-60 | 1 |
| Marital status | Single | 2 |
|  | Married | 11 |
|  | Divorced | 1 |
| Family composition | 1-2 | 5 |
|  | 3-4 | 4 |
|  | 5+ | 5 |
| Profession | Doctors | 7 |
|  | Nurses | 5 |
|  | Admin | 2 |
| Working experience | 1-5 | 6 |
|  | 6-10 | 3 |
|  | 11+ | 5 |
| Managing COVID-19 patients | Yes | 13 |
|  | No | 1 |
| Vaccinated | Yes | 9 |
|  | No | 5 |
| Views about Vaccination | It helps | 8 |
|  | It does not help | 3 |
|  | Not sure | 2 |
| Trained on COVID-19 | Yes | 9 |
|  | No | 5 |
| Infected with COVID-19 | Yes | 6 |

### Initial experience of FLHW in dealing with COVID-19 pandemic

*Normal*. Participants had a variety of responses about their initial experience responding to the COVID-19 pandemic. Some took it as a normal part of the ups and downs of life and there were no prolonged drastic changes in attitude or extreme thoughts and emotions when they first heard about the pandemic.

> *“At the beginning, there were times I felt that I was infected, but as days went on, I got used to it. Sometimes you feel like you are sick but you move on and nothing happened. For the time being, COVID-19 is not a big threat anymore, we are used to it”* (Participant 7, medical personnel)

*Uncertainty.* Others were uncertain about the seriousness of the pandemic leaving them unsure on how to approach the situation and how its progression would affect their professional and personal lives. There was information flooding about the pandemic from media to religious views and political denial that increased the uncertainty of the progression of COVID-19 and uncertainty of how to handle it.

> *“It was discouraging because we were unsure about what we were doing.”* (Participant 7, medical personnel)

It was difficult for some participants to come to terms with the reality of the pandemic instead responded by denying its existence and continuing their activities as if no changes were happening around them. Some felt that denying the existence of COVID-19 would decrease the emotional burden they would have to deal with.

### Impact of COVID-19 on Front-line Health Workers

#### Psychological impact

##### Fear

Participants reflected on their fear of getting infected and infecting others especially family as they witnessed increased patient deaths including close relatives and acquaintances. Long working hours that exposed them to the rising toll made them fear that they would also die from COVID-19 making others despair.

> *“Fear was too much because we lost many relatives”* (participant 4, nurse)

> *“There was a time I believed that I would die from covid 19, so I had to spend my time to clear with God and other people” (be in harmony with God and other people)* (Participant, medical Doctor)

> *“Many times, when I was transporting patients, I didn’t observe the medical rules because I knew I would die anytime* (Participant 3, Ambulance Driver)

##### Stress

The sudden change in the working environment to accommodate the pandemic meant long labor and emotionally intensive working hours. Some participants would work double shifts to accommodate for the limited human resource and new protective measures were strict and uncomfortable for the health workers.

> *‘In the beginning, it was very stressful, even putting on the mask and other protective gear throughout the day was a punishment to me”* (Participant 6, Nurse)

> *“It was very challenging for me because you were carrying patients from the emergency department to the isolation ward. You can carry about 15 patients during the day time then at night you find among them you carry10 from the isolation ward to the mortuary”. So, it was very challenging for me”* (Participant 7, Pathology supporting staff)

### Stigma/Discrimination

There was fear of being infected among staff and from the community. FLHW felt alone and unable to be assisted when needed because other people would avoid getting in contact with them.

> *“Outside the office and even in the workplace, people were afraid of me, because as a driver, I used to transport COVID-19 patients, and it was very difficult to convince the people that I was not infected. Sometimes I afraid to go outside shopping because people were avoiding me, when they saw me they moved away, I once cried about this”* (Participant 3, Ambulance Driver).

### Impact on Professionalism

Several significant themes emerged highlighting the impact of COVID-19 on professional and ethical obligations. Some health workers wanted to quit their job due to the strenuous workload and emotional burden while other absence themselves from work and other were running from attending patients due to fear. Others questioned whether their career choice was appropriate and their ability to handle the obligations expected of them at the time.

> *“If I could find another job I could quit this one.”* (Participant 12, nurse)

### Absenteeism

There was an increased number of health workers who avoided signing up to work in isolation wards and increased absenteeism for those who were there.

> *“Some staff who used to cheat, they wrote letters saying that “my health is not good, I can’t come to work” so some staff were dodging the work”* (participant 4, nurse)

### Avoiding patients

Some health workers were adamant about receiving patients due to fear of infection and the limited capacity of the hospital to accommodate patients. There was also concern about the availability of utilities to facilitate the treatment of patients.

> *“At the beginning, we were reluctant to receive COVID-19 patients due to fear of infection and capacity of the hospital to manage the patients”* (participant 2, administrative secretary)

### Vaccination refusal

While some health workers were opposed to receiving vaccination, others felt that it did not provide adequate protection. Some health workers believed the rapid changing of the virus made the available vaccines less viable and it was better to go without them.

### Communication with Relatives

Health workers had to observe social distancing rules while talking to relatives of patients which proved difficult to adhere to in the beginning of the pandemic prompting frequent reminders to relatives. There was also a need for constant updates on the patients’ progress and a lot of time was used in educating relatives even during funerals.

> *“We used to escort the dead body and provide education to the family members during the funeral service-this was stressful additional work”* (Participant 3, a nurse)

### Conflict in Work Place

The ever-changing protocols and high-stress environment increased conflict between FLHW and their seniors as they tried to navigate the burden of care and other professional responsibilities. Communication was brief and FLHW would sometimes feel frustrated due to limited resourceful information that would help them provide care more efficiently as one participant noted;

> *“Due to the nature of the situation, there was a time when we had a lot of conflicts with our bosses because there was no time to discuss issues, you just have to receive the order and implement. Everyone was under stress” (Participant 2, administrative nurse)*.

### Perception of Caring for COVID-19 Patients

Participants described the experience of caring for COVID-19 patients to be a “***Life and Death calling***” or “***an offering***.” These words depict the intensity of their experiences and the heavy sense of duty that they had as healthcare providers during this time. The sense of urgency and always being faced with high-stakes and constant decision-making in saving the lives of patients. As one participant said;

> *“Because we were sacrificing ourselves, you just work “kama sadaka “(as an offering), a medical doctor”*.

### Uncertainty with Appropriate Treatment - Trial and Error

Treatment protocols were changed multiple times during the pandemic. There were a lot of suggestions about management and care from different places around the world which created frustration as to which methods were more effective. However, even with these suggested management had to be adjusted to keep up with the changing symptom presentations of the virus.

> *“There was no a system in place but people were adjusting according to the challenges”* (Participant 3, Nurse)

> *“It was discouraging because we were unsure about what we were doing and how to handle patients, so it was like trial and error.”* (Participant 7, medical personnel)

### Uncertainty in Handling Patients

Apart from the ever-changing management, FLHW also experienced uncertainty in handling patients due to feeling unprepared and not knowing what they were dealing with. For the majority of participants, it was their first time handling a pandemic therefore they had no prior experience to reference from.

> *‘I have never seen that thing, I don’t know’* (participant 2, administrative secretary)

### Difficulty Adapting to New Treatment Modalities and Policies

The rapid changes in the management of patients made it difficult for staff to keep up and the continuous need to learn and implement new techniques meant a steep learning curve. Before getting comfortable with the existing techniques, they would find themselves thrown off balance again to keep up with the ever-changing treatment options. Not enough coherent training was provided which made care of patients inconsistent as workers tried to keep up with the changing guideline and policies.

> *“There was no system in place but people were adjusting according to the challenges and new directives.”* (Participant 13, medical doctor)

### Difficulty Adapting to Preventative Measures

Some participants reported a hard time dealing with the specific additional preventive measures that were necessary to prevent the increase in the outbreak and infection of COVID-19. The requirement to wear personal protective equipment (PPE) all the time and strictly follow disinfection measures, was an additional challenge on top of their busy schedules and left them feeling physically uncomfortable.

> *“Some health care workers couldn’t properly observe medical procedures due to fear of being infected (Participant no 5, medical doctor)*

### Failure to Observe Proper Procedures Due to Fear of Infection

Some of the FLHW workers had no idea what kind of preparation they needed to make for a pandemic, which only added to their feeling of being inadequate. The unpredictability of the disease and the novelty of experience from FLHW meant that they were learning about everything in real time.

> *“At the beginning, we were reluctant to receive COVID-19 patients due to fear of infection and capacity of the hospital to manage the patients”* (Participant 2, administrative secretary)

> *“Some HCW couldn’t observe proper medical procedures due to fear of being infected* (Participant 9, a nurse)

### Dilemma in Self-Care and Patient Care

Participants had very limited time for self-care due to the long working hours, sometimes taking up double shifts. They had difficulty prioritizing their health and well-being without feeling guilty about their professional duty to care for patients. This exposed them to conflict in work-life balance further enhancing their stress. There were a lot of informal messages about preventative measures that some participants also found themselves using to prevent getting infected.

> *“In those days, after learning the details about COVID-19 and its treatments were not formalized—everyone was doing as they wished—so I started using ginger, oranges and other local herbs.”* (Participant 8, Nurse).

### Coping strategies

Participants described different strategies to manage the stress of the pandemic and maintain resilience. Strategies varied from acceptance, resorting to religious faith, seeking comfort from family, and resting, to avoidance strategy such as use of substance like alcohol and cannabis, denial, and, isolation.

Some participants were able to accept the presence of the pandemic. They were ready to accept the reality and adapt to the changes required to face the adversity they were in. for some it wasn’t easy to face the reality of the situation or the need for emotional support for health workers so they resorted to keeping themselves busy with activities so they would not have to think about the pandemic while some felt they had no one to turn to when they needed support, feeling very lonely and isolated from the world.

> *“I needed it (emotional support) but I didn’t know I needed it; I was hiding behind other things.”*

> *‘My mother was my comforter, so after her death, I became lonely, I had no one I can go to and explain my problems, “no” so I had very lonely life”* (Participant 2, administrative secretary)

### Faith and prayer

Some found solace in reading Holy books and increasing their faith-based activities as a way to gain clarity of the situation and maintain hope amid the chaos.

> *“I am a devoted believer, so I often read the word of God, and it was very comforting, when I got home, I first rest for an hour and I had quiet time at my house to read Bible verses to comfort myself.”* (Participant 8, Nurse)

> *“There was a time I believed that I would die from COVID-19, so I had to spend my time to clear with God and other people, I wanted to be in harmony with God and other people, medical doctor”* (Participant 7, a Doctor)

### Family comfort

Having a supportive family environment was a source of comfort for some health workers that helped improve their ability to cope.

> *“if I was with my husband, I would talk to him about how to wear a mask and if you tell him something, he would listen.”* (Participant 12, a nurse)

> *‘My mother was my comforter, so after her death, I became lonely, I had no one I could go to and explain my problems, “no” so I had a very lonely life”* (Participant 2, administrative secretary)

### Resting and Use of Recreational Substances

When they were off duty, some made sure to get as much rest as possible to recuperate from the stress of the shifts. Some health workers use recreational substances to help them feel relaxed and aid in sleeping.

> *“like others, you find that you are tired, so to relax, you drink two beers, you rest and sleep.”* (Participant 14, medical doctor)

## Discussion

This study explored the lived experiences impact and coping strategies of frontline health workers in Kilimanjaro region during the COVID-19 pandemic. Four key themes emerged from in-depth interviews with healthcare workers namely: 1. Initial response to pandemic, 2. impact of professionalism, 3. Impact of pandemic on care and treatment and 4. Coping strategies with pandemic.

### Initial response to pandemic

Responses ranged from denial and uncertainty to experiencing no drastic change in attitude or emotional response reflecting the complexities and range of human reactions to adverse situations. An integrative review of previous studies showed that individuals who saw the pandemic as part of life’s ups and downs were likely to have positive outcomes, more likely to have higher levels of resilience and ultimately better-coping mechanisms than those who didn’t [5]. Limited knowledge and experience with COVID-19 expressed by participants align with findings of uncertainty prompting early treatment and investigation while increasing the risk of overexertion and potential misuse of resources [6]. This was echoed by participants description of rapidly changing management protocols. These findings show the need for incorporating updated and ongoing psychological readiness and safety procedures for FLHWs.

### Impact of Pandemic on Professionalism Care and Treatment

The COVID-19 pandemic presented many ethical challenges to FLHW, ranging from allocating scarce resources, to balancing a duty of care with self-preservation, and implementing visitation restrictions. In our study some health workers thought of quitting their job due fear of death, the strenuous workload and emotional burden while others questioned their career and some were avoiding patients to protect their life resonating to studies from other parts of the world [7, 8, 2,9].

Increased absenteeism in our findings was attributed to the fear of getting an infection and lack of support as pointed out by other researchers [10] and increased distrust among colleagues [11]. Additionally, fluctuations in the practices of managing patients posed serious challenges to FLHW as they grappled to adapt to the everchanging guidelines [12,13,6,1]. As a result of the influx of conflicting information regarding the spread of COVID-19, some FLHW were drawn to use traditional/ non-scientific methods to prevent themselves from infection [14].

Given these findings, timely, clear and evidence-based feedback should be given to FLHW to avoid misinformation and underlying uncertainty. Ensuring resource availability and manageable working hours is key in reducing absenteeism, fear and moral distress. Future preparedness should include collaborative approach to decision-making wherever possible and resilience training to maintain professionalism and quality of care during emergencies.

### Coping strategies of FLHW

COVID-19 had a huge impact on FLHW necessitating adoption of different coping strategies. A wide range of strategies both healthy and unhealthy, were shared by study’s participants as a way of responding to the challenges brought about by the pandemic. Acceptance helped participant adapt to the new “normal” aligning with findings that it fosters resilience by focusing on controllably aspects [2]. Prayer and Faith provided comfort and clarity reflecting on importance of spiritual and religious practices in enhancing psychological well-being during turbulent times [15]. Support form close family such as providing room to share challenges offered safety and moral support while buffering against stress and promoting resilience [16] while resting during off-hours enabled FLHWs decrease work related stress [17].

On the other hand, some participants relied on substances to as a coping strategy but this is known to have potential negative outcomes [18]. Other participants used isolation and avoidance as a coping strategy although they have been associated with increased stress and anxiety [19,20]. Denial was experienced by some of the participants initially and research supports that it is usually a strategy that is used short-term as individuals develop other long-term coping abilities [4,21].

It is essential to have to provide targeted mental health interventions to address unhealthy coping mechanisms such as substance use, isolation and prolonged denial which are known to increase distress. Integrating this from policy level will ease adaption into current health systems and encourage practice for self-care and help seeking behavior in FLHW.

## Lessons learned

Throughout the in-depth interviews, several key lessons emerged as they reflected on their experiences.

### Strengthening protective measures

FLHW recognized the importance of following protective measures to reduce the risk of infecting themselves and others. The pandemic has helped them to better understand their role in preventing infections while still being able to provide proper care to patients. This points to the need for continued facility level support to ensure access to protective equipment, training on infection and prevention control beyond pandemics. It also highlights importance of policies that have these practices embedded into routine health systems operations.

### Resilience, confidence, and patient centered care

Participants shared how the experience of working during the pandemic has made them stronger and better able to face the adversities of work and life in general. They also learned to be more compassionate about patients,

Health systems should consider structured psychosocial support and resilience building initiatives to sustain current growth and encourage preparedness for future pandemics.

> *“COVID-19 has helped me to be more compassionate to patients because I witnessed a lot of deaths due to COVID-19 at that time, it has taught me that everyone can die at any time, It has increased my love for others”* Participant 13, medical doctor.

### Study strengths and limitations

One of the strengths of this qualitative study is the depth of viewpoints regarding the impact of the pandemic. We tried to include participants from different departments who were in direct contact with patients and their relatives including ambulance drivers, morticians, and secretaries.

We acknowledge that while the overall participant sample was purposive, the duration between our study and the last wave of the pandemic was long. Time and continued life experiences may have impacted the richness of the data collected.

## Conclusion

This is the first qualitative study to explore the lived experiences and coping strategies of the FLHW in dealing with the COVID-19 outbreak in Tanzania. Results show that the COVID-19 pandemic has challenged FLHW, with multiple contextual factors such as limited human and material resources, and, rapid changes of treatment protocols impacting their experiences and needs. Participants experienced fear, stress, and stigma that emanated from the pandemic and these emotions affected their employment and interpersonal relationships. Healthy coping mechanisms such as accepting one’s fate and trusting in the Lord were used as well as unhealthy strategies such as substance abuse and social ostracism. These findings can be used to guide the design integrating and comprehensive psycho-social support and participatory management to mitigate the challenges of pandemics in similar settings in the future.

COVID-19 pandemic has undoubtedly changed the care practices, perception, and emotional strength of FLHW. Mandatory use of protective measures increased awareness and compliance with the recommended infection control measures. The pandemic itself has boosted the staff’s critical and coping skills in working through the challenges brought by disease, thus developing the healthcare workers’ readiness for other future adversities. However, the pandemic has also increased healthcare workers’ level of compassion and gratitude as compared to pre-pandemic times. All of these changes speak to the deeply transformative effects that the pandemic has had on the professional and individual existence of FLHW. Since the study highlights COVID- 19 had an impact on professionalism and patients, we also recommend health care facilities to have pandemic prepared plans and regularly conduct pandemic preparedness training with emphasis on mental health among FLHCWs.

## Data Availability

The minimal data set set supporting the findings f this study are available in the manuscript. Full qualitative manuscripts cannot be made available due to ethical restrictions however anonymized excerpts are provided.

## Acknowledgements

The authors would like to express their sincere gratitude to the KCMC frontline health care workers who took their time to share their experienced during an incredibly challenging time. We also acknowledge the support of the hospital for facilitating accessibility to participants. Special thanks to research assistants and transcribers for their valuable contribution throughout the study.

